# Long-term Occupational Exhaust Fumes Exposure and Delayed Cognitive Impairment in Older U.S. Adults: A Cross-sectional Study in U.S

**DOI:** 10.1101/2025.06.16.25329589

**Authors:** Lili Liang, Perry E Sheffield, Rose Saint Fleur-Calixte, Tianxu Xia, Spencer Xinyi Zhang, Jenny J Lin

## Abstract

**Background:** Occupational exposure to exhaust fumes, containing neurotoxic particulate matter and polycyclic aromatic hydrocarbons (PAHs), is associated with cardiopulmonary diseases, but its cognitive effects in aging workers remain insufficiently studied. Given increasing occupational longevity, understanding these risks is critical for dementia prevention.

**Methods:** We analyzed data from 1,110 adults aged 60 years and older in the 2011–2012 National Health and Nutrition Examination Survey (NHANES), comparing cognitive performance between exposed (24%) and unexposed groups. Cognitive function was assessed using the Consortium to Establish a Registry for Alzheimer’s Disease Word List Learning Test (CERAD-WL), Animal Fluency Test (AFT), and Digit Symbol Substitution Test (DSST). Distributed lag nonlinear models (DLNMs) evaluated non-linear and time-lagged effects of exposure duration.

**Results:** Exposed workers were predominantly male (81.6% vs. 41.8%), had lower educational attainment (31.2% vs. 24.4% with less than high school education), and exhibited higher rates of smoking (65.0% vs. 48.6%) and excessive alcohol use (15.7% vs. 7.0%). Occupational exposure was associated with significant cognitive impairments in delayed memory (odds ratio [OR] = 2.55, 95% confidence interval [CI]: 1.61–4.05), verbal fluency (OR 2.41, 1.48–3.94), and processing speed (OR 1.95, 1.19–3.18). The DLNM analyses revealed a biphasic response: minimal effects at <20 years of exposure, but major declines after 30 years, with a 15–25-year latency period.

**Conclusion:** Prolonged occupational exhaust fume exposure is associated with domain-specific cognitive decline, particularly affecting memory and executive function. The dose-response relationship underscores cumulative neurotoxicity, emphasizing the need for targeted protections for high-exposure workers.

## 1. Introduction

Exhaust fumes represent a major and complex source of environmental exposure, consisting of fine particulate matter (PM2.5), polycyclic aromatic hydrocarbons (PAHs), volatile organic compounds (VOCs), nitrogen oxides (NOx), metals, and other reactive species capable of penetrating deep into the respiratory system and even crossing the blood–brain barrier (Hänninen et al., 2025). A growing body of epidemiological evidence has linked chronic exposure to ambient air pollution with cognitive decline, dementia, and Alzheimer’s disease (Park et al., 2022; Parra et al., 2022; Semmens et al., 2023). Observational studies focusing specifically on airborne pollutants such as PM2.5 and combustion byproducts further suggest that long-term exposure accelerates cognitive dysfunction in older adults (Shaffer et al., 2021).

The mechanisms underlying these associations are thought to involve both direct and indirect pathways. Ultrafine particles may reach the brain through the olfactory bulb, while systemic inflammation and oxidative stress may indirectly impair neural function (Yokota et al., 2015). Animal models have demonstrated acute and subchronic neuroinflammation, oxidative injury, and markers of neurodegeneration after exposure to exhaust fumes (Coburn et al., 2018; Milani et al., 2020). Consistent with these biological findings, behavioral experiments reveal impairments in learning, memory, and spatial cognition in exposed rodents (Li et al., 2020; Garrick et al., 2023). Human studies provide parallel evidence: reductions in school bus exhaust exposure improved students’ performance, while short-term exposure to combustion-related pollutants among commuters and office workers was associated with diminished attention, processing speed, and executive function (Pedde et al., 2024; Mallach et al., 2023; Yang et al., 2024). Collectively, these findings suggest that both acute and chronic exposure to exhaust fumes may impair cognitive performance.

Despite this evidence, the impacts of occupational exhaust fume exposure remain underexplored. Beyond traffic exhaust, workers in transportation, construction, firefighting, aviation, and personal care industries may be exposed to a real-world mixture of pollutants that includes not only combustion products but also PFAS, heavy metals, and solvent vapors. These pollutants may act synergistically with workplace stressors such as noise and psychosocial strain, thereby compounding risks of systemic inflammation, hypertension, and metabolic syndrome, all of which are linked to cognitive decline (Wright et al., 2014; Costache et al., 2023). However, the long-term neurotoxic effects of occupational exposure, including latency periods between exposure and the onset of cognitive decline, remain poorly understood, limiting the development of preventive strategies for aging workforces.

To address this gap, the present study uses data from the 2011–2012 National Health and Nutrition Examination Survey (NHANES) to investigate the relationship between occupational exhaust fume exposure and cognitive function in adults aged 60 years and older. Using a novel application of Distributed Lag Nonlinear Models (DLNM), this study aims to characterize latent risk patterns and nonlinear dose–response relationships. Findings are expected to strengthen the evidence base on occupational environmental exposures and neurocognitive health, providing critical insights for workplace risk assessment, cognitive health monitoring, and the development of targeted preventive strategies.

## 2. Materials and methods

### 2.1 Study population

We used NHANES data collected in the 2011–2012 cycle. This analysis focused on older adults aged 60 years and above to explore the association between occupational exposure to exhaust fumes and cognitive function. Participants were eligible for inclusion if they had completed all cognitive function assessments and had complete data on occupational exposure to exhaust fumes, demographic characteristics, and relevant covariates (e.g., educational attainment, household income, race/ethnicity, smoking and alcohol use). Following the inclusion criteria outlined in Figure 1, a total of 1110 adults were included in the study.

**Figure 1.**
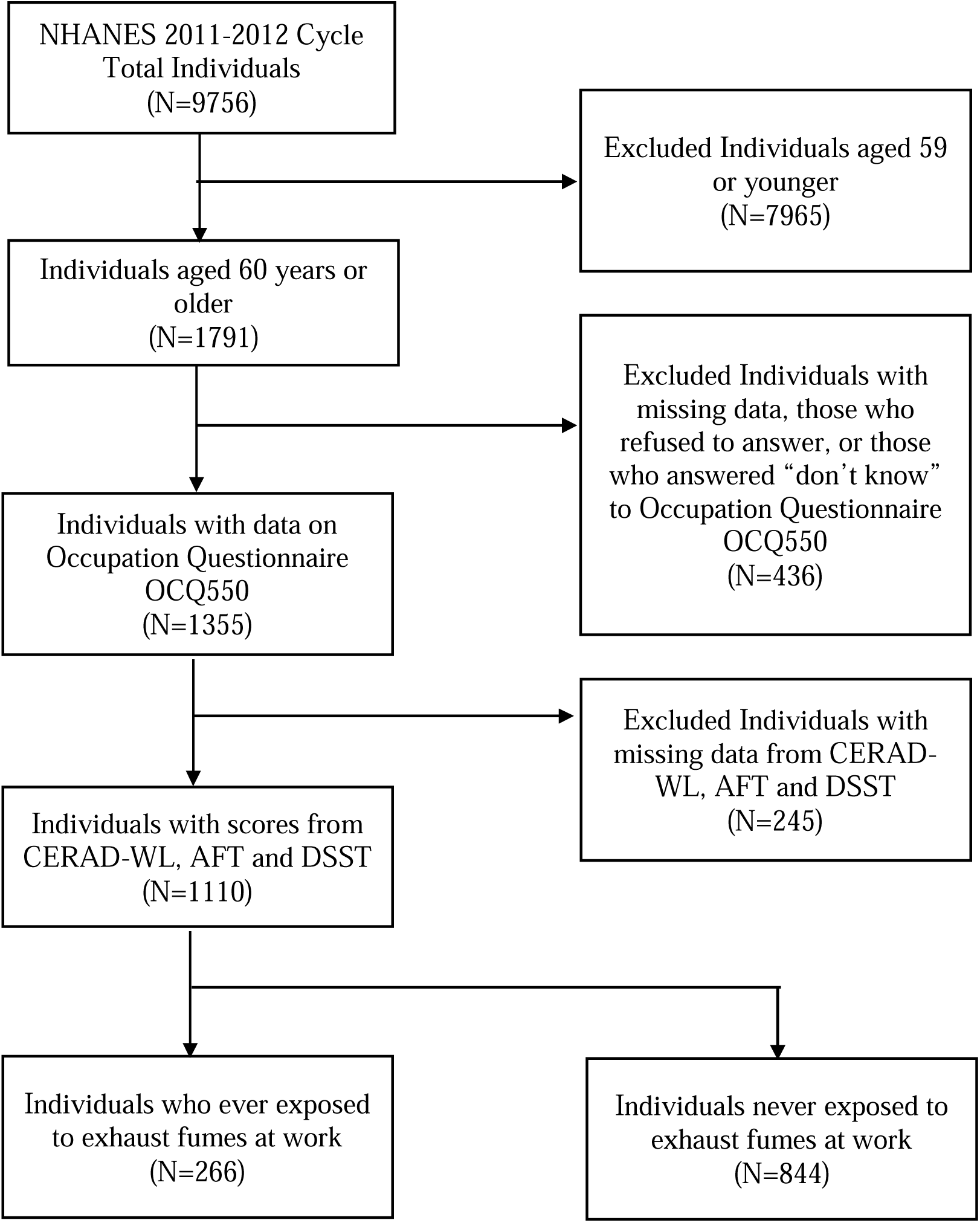
Flowchart for Selecting Eligible Population. Notes: NHANES, National Health and Nutrition Examination Survey. OCQ: occupation questionnaire. CERAD-WL, Consortium to Establish a Registry for Alzheimer’s Disease Word List Learning Recall Test. AFT, Animal Fluency Test. DSST, Digit Symbol Substitution Test.

### 2.2 Measurement of occupational exposure history

The Occupation Questionnaire section of the NHANES 2011-2012 study includes interview data regarding employment and work environment-related variables. A subset of individuals aged 16-79 were asked about their lifetime history of exposure to mineral and organic dusts, gases, fumes, and vapors in the workplace, specifically, participants were first asked whether they had ever been exposed to exhaust fumes at work (OCQ550), defined as breathing in fumes or having a lingering smell on their clothes, skin, or hair. These exhaust fumes could originate from sources such as trucks, buses, heavy machinery, and diesel engines in any job. Additionally, participants who answered ‘yes’ were then asked to report the total number of years they had been exposed to exhaust fumes at work from all jobs (OCQ560). OCQ550 (binary, exposed or unexposed) was used to categorize participants. OCQ560 (years of exposure) was treated as a continuous variable.

### 2.3 Cognitive function Assessment

Participants aged 60 years and older were eligible to cognitive function Assessment. Cognitive function was evaluated using three standardized tests: Consortium to Establish a Registry for Alzheimer’s Disease Word List Learning Test (CERAD-WL): This test measures both immediate and delayed recall abilities for new verbal information (Morris et al., 1989). This assessment includes three sequential learning trials followed by a delayed recall test. In each learning trial, participants are asked to read aloud a list of 10 unrelated words. Immediately after hearing the list, participants are instructed to recall and recite as many words as they can remember. The delayed recall test takes place roughly 10 minutes after the start of the initial word learning trials. Each trial allows for a maximum score of 10, with a total possible score of 40, which includes the combined scores from all three trials plus the delayed recall.

Animal Fluency Test (AFT): This test evaluates verbal category fluency, which is an aspect of executive function, alongside other cognitive functions like semantic memory and processing speed (Clark et al., 2009). Participants are required to name as many different animals as they can within a one-minute period, earning one point for each correctly named animal.

Digit Symbol Substitution Test (DSST): This test serves as a measure of working memory, assessing skills such as processing speed, sustained attention, and short-term memory (Wechsler et al., 1997). This test involves a paper form where participants are given a key displaying nine numbers, each paired with a unique symbol. Over the course of two minutes, participants must accurately copy the corresponding symbols into 133 boxes aligned with the numbers.

According to the guidelines of the Uniform Data Set (UDS) (Dodge et al., 2020), cognitive impairment is defined as an adjusted Z-score ≤ -1.5 standard deviations (SD) (based on age- and education-corrected norms), with cognitive performance dichotomized as “impaired” or “unimpaired” to identify cognitive impairment.

### 2.4 Covariates

Demographic variables included age (either 60–69 or 70–79 years), gender (male, female), race or ethnicity (Hispanic, non-Hispanic White, non-Hispanic Black, non-Hispanic Asian and other race), and educational attainment (< HSG: less than 12th grade, including 12th grade with no diploma; HSG: high school graduate or GED; >HS: some college or associate degree, and college graduate or higher). Health-related variables included body mass index (BMI), smoking history, alcohol use, high blood pressure, abnormal cholesterol, heart disease, depression, sleep problems, and hyperglycemia. BMI (kg/m²) was calculated using height and weight measured by trained staff. Obesity was defined as a BMI ≥30 kg/m². Smoking history was divided into two groups: those who smoked at least 100 cigarettes in their life and those who did not (Zhu et al., 2023). Excessive alcohol use was defined as more than 350 g/week for females or more than 420 g/week for males (Kalligeros et al., 2024). High blood pressure was defined by a measured blood pressure of 140/90 mmHg or higher or if the person reported taking prescription for hypertension. Dyslipidemia was defined by triglyceride levels of 1.70 mmol/L (150 mg/dL) or higher, low-density lipoprotein cholesterol of 3.4 mmol/L (130 mg/dL) or higher, or high-density lipoprotein cholesterol less than 1.0 mmol/L (40 mg/dL) in men and less than 1.3 mmol/L (50 mg/dL) in women. History of cardiovascular diseases (CVD) was reported by participants and included history of heart failure, coronary heart disease, angina, heart attack or stroke. Depression was measured using the PHQ-9 questionnaire, with scores of 10 or higher indicating depression. Sleep problems were identified if participants said they slept 1–4 hours per day or said “yes” to the questions, “Did a doctor ever say you had trouble sleeping?” or “Did a doctor ever say you had a sleep disorder?” (Lee et al., 2021) Hyperglycemia was defined by fasting glucose of 5.6 mmol/L (100 mg/dL) or higher, hemoglobin A1c of 5.7% (39 mmol/mol) or higher, or if the person said they had type 2 diabetes or were being treated for it.

### 2.5 Statistical analysis

Analyses accounted for the complex sampling design of NHANES by incorporating recommended sampling weights, strata, and primary sampling units to ensure representative results, performed using SAS software (version 9.4, SAS Institute Inc., Cary, NC) and R (v4.4.1).

Descriptive statistics were calculated for demographic and health-related variables. Differences between exposed and unexposed groups were assessed using Chi-square tests for categorical variables. The association between occupational exhaust fume exposure (binary) and cognitive impairment (binary) was evaluated using weighted, multivariable logistic regression models, adjusted for potential confounders including age, gender, race/ethnicity, educational attainment, and health behaviors/comorbidities.

Standard regression models are limited in their ability to capture potential nonlinear exposure-response relationships and delayed (lag) effects. We therefore employed Distributed Lag Nonlinear Models (DLNMs) to investigate the complex relationships between years of exposure (OCQ560, continuous variable) and cognitive function scores (continuous variables). A cross-basis function was constructed using natural splines (df = 3), with a maximum lag duration of 10 years. Effect estimates (β coefficients with 95% CIs) were visualized with the statistical significance set at a two-tailed α = 0.05.

To explore inter-domain relationships, we calculated Spearman correlation coefficients among three cognitive tests (CERAD-WL Delayed Recall, AFT and DSST). A Sankey diagram was used to visualize the distribution of multi-domain cognitive impairment patterns across exposure groups.

Missing data were handled using an available-case analysis approach. Every statistical model was fitted using all participants with complete data for the variables in that specific analysis. No data imputation was performed.

### 2.6 Ethical approval

This study was approved by the Institutional Review Boards of the Icahn School of Medicine at Mount Sinai.

## 3. Results

### 3.1 Participant Characteristics

Based on Table 1, a total of 1,110 participants aged ≥60 years were included, with 266 (24.0%) reporting occupational exhaust fume exposure. Exposed individuals were predominantly male (81.6% vs. 41.8%, P < 0.0001), had lower educational attainment (31.2% vs. 24.4%, P = 0.017), and higher rates of smoking (65.0% vs. 48.6%, P < 0.0001) and excessive alcohol use (15.7% vs. 7.0%, P = 0.001) compared to the unexposed group. Additionally, we compared baseline characteristics between participants who completed all cognitive tests and those who did not, and the comparison revealed no substantial differences, suggesting a low risk of attrition bias (Supplementary Table s1).

**Table 1.** Baseline Characteristics of Participants by Exposure Status (n=1110). Notes: Education Level: < HSG: less than 12th grade, including 12th grade with no diploma; HSG: high school graduate or GED; >HS: some college or associate degree, and college graduate or higher). CVD, Cardiovascular Diseases. PHQ-9, Patient Health Questionnaire-9.

|  | **No.**<br>**(%)** | **Exposed**<br>**group**<br>**No. (%)** | **Never Exposed**<br>**group**<br>**No. (%)** | ***P***<br>***Value*** |
| --- | --- | --- | --- | --- |
| **Total** | **1110**<br>**(100.0)** | **266**<br>**(24.0)** | **844**<br>**(76.0)** |  |
| **Age (years)** |  |  |  | ***0.576*** |
| 60-69 | 727<br>(65.5) | 178<br>(16.1) | 549<br>(49.4) |  |
| 70-79 | 383<br>(34.5) | 88<br>(7.9) | 295<br>(26.6) |  |
| **Gender** |  |  |  | ***0.0001*** |
| Male | 570<br>(51.4) | 217<br>(19.6) | 353<br>(31.8) |  |
| Female | 540<br>(48.6) | 49<br>(4.4) | 491<br>(44.2) |  |
| **Race/Ethnicity** |  |  |  | ***0.019*** |
| Mexican American and Other<br>Hispanic | 214<br>(19.3) | 51<br>(4.6) | 163<br>(14.7) |  |
| Non-Hispanic White | 435<br>(39.2) | 113<br>(10.2) | 322<br>(29.0) |  |
| Non-Hispanic Black | 347<br>(31.2) | 88<br>(7.9) | 259<br>(23.3) |  |
| Non-Hispanic Asian and<br>Other Race | 114<br>(10.3) | 14<br>(1.3) | 100<br>(9.0) |  |
| **Education Level** |  |  |  | ***0.017*** |
| < HS | 290<br>(26.1) | 84<br>(7.5) | 206<br>(18.6) |  |
| HSG | 245<br>(22.1) | 65<br>(5.9) | 180<br>(16.2) |  |
| > HS | 575<br>(51.8) | 118<br>(10.6) | 457<br>(41.2) |  |
| **Smoking history (Ever smoke)** |  |  |  | **0.0001** |
| >100 cigarettes | 583<br>(52.5) | 173<br>(15.6) | 410<br>(36.9) |  |
| < 100 cigarettes | 527<br>(47.5) | 93<br>(8.4) | 434<br>(39.1) |  |
| **Excessive Alcohol Use<sup>†</sup>** |  |  |  | **0.001** |
| Yes | 59<br>(9.3) | 26<br>(4.1) | 33<br>(5.2) |  |
| No | 576<br>(90.7) | 140<br>(22.0) | 436<br>(68.7) |  |
| **Obesity** |  |  |  | **0.107** |
| Yes | 466<br>(42.0) | 123<br>(11.1) | 343<br>(30.9) |  |
| No | 644<br>(58.0) | 143<br>(12.9) | 501<br>(45.1) |  |
| **Hypertension** |  |  |  | **0.555** |
| Yes | 655<br>(59.0) | 161<br>(14.5) | 493<br>(44.5) |  |
| No | 455<br>(41.0) | 105<br>(9.5) | 350<br>(31.5) |  |
| **Dyslipidemia\*** |  |  |  | **0.290** |
| Yes | 645<br>(62.6) | 150<br>(14.5) | 495<br>(48.0) |  |
| No | 386<br>(37.4) | 101<br>(9.8) | 285<br>(27.7) |  |
| **Hyperglycemia<sup>§</sup>** |  |  |  | **0.640** |
| Yes | 759<br>(71.3) | 187<br>(17.6) | 572<br>(53.7) |  |
| No | 305<br>(28.7) | 71<br>(6.7) | 234<br>(22.0) |  |
| **History of CVD** |  |  |  | *0.098* |
| Yes | 204<br>(18.4) | 58<br>(5.2) | 146<br>(13.2) |  |
| No | 906<br>(81.6) | 208<br>(18.7) | 698<br>(62.9) |  |
| **Depression (PHQ-9 score)** |  |  |  | *0.215* |
| Yes ($\geq 10$) | 105<br>(9.5) | 20<br>(1.8) | 85<br>(7.7) | |
| No ($< 10$) | 1005<br>(90.5) | 246<br>(22.1) | 759<br>(68.4) | |
| **Have Sleep Problems** |  |  |  | *0.860* |
| Yes | 379<br>(34.3) | 92<br>(8.3) | 287<br>(26.0) |  |
| No | 727<br>(65.7) | 173<br>(15.6) | 554<br>(50.1) |  |
| ***§ Missing data: Excessive Alcohol Use (475), Dyslipidemia (79), hyperglycemia (46) |  |  |  |  |

### 3.2 Cognitive Function and Exposure Associations

Occupational exhaust fume exposure was associated with significant cognitive decline across all tests (Figure 2). The strongest negative effects were observed for delayed memory (CERAD-WL delayed recall: OR = 2.55, 95% CI: 1.61–4.05), verbal fluency (AFT: OR = 2.41, 95% CI: 1.48– 3.94), and working memory (DSST: OR = 1.95, 95% CI: 1.19-3.18), while immediate recall showed no significant association.

**Figure 2.**
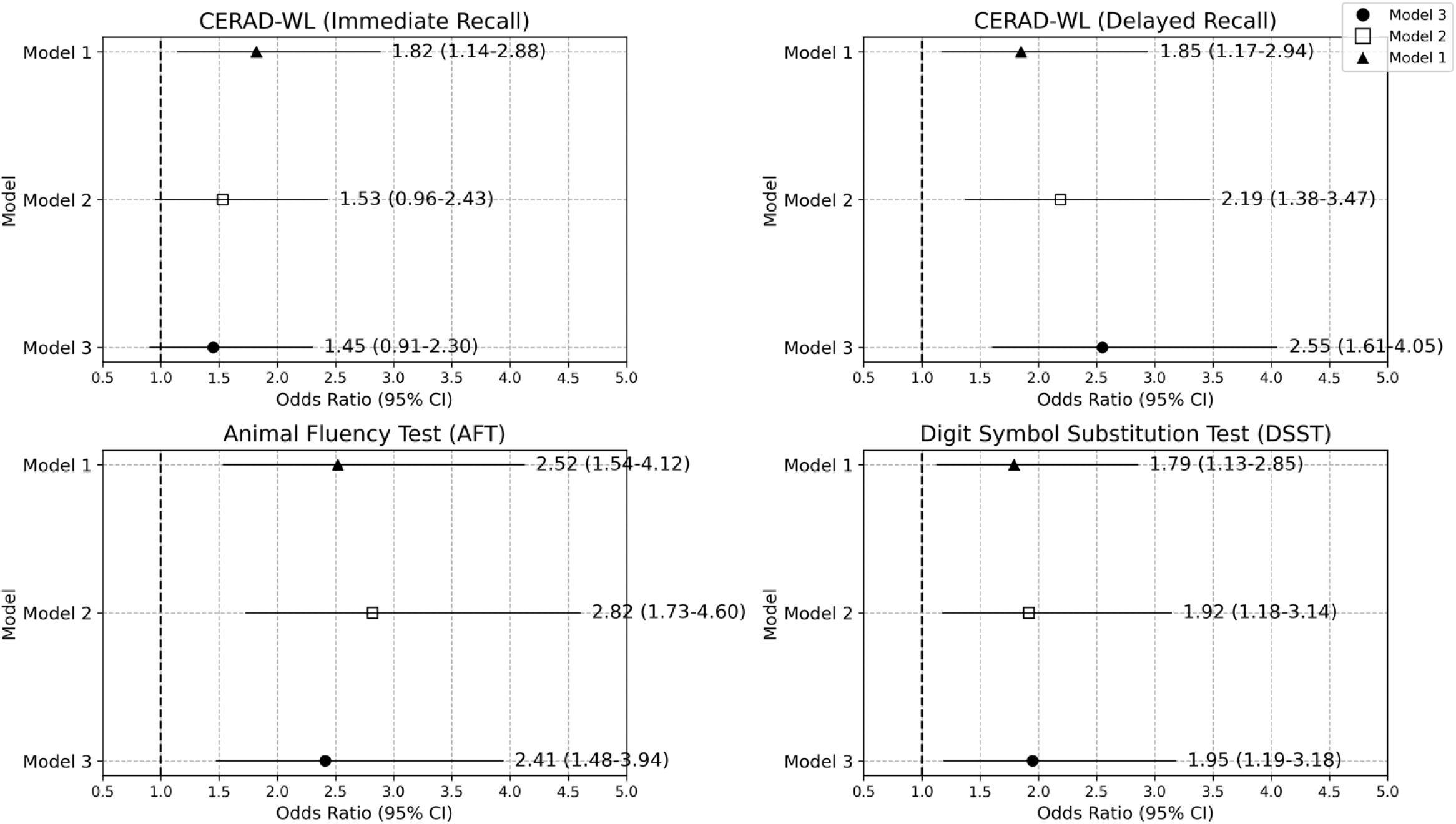
Forest Plot of Weighted Multivariable Survey Logistic Regression Analyses on the Association Between Occupational Exhaust Fumes Exposure and Cognitive Decline (n = 1,110). Adjusted for: Model 1, no adjustment; Model 2, age, gender, and educational attainment; Model 3, age, gender, educational attainment, race, history of CVD, depression and diabetes. Notes: CVD, cardiovascular diseases. CERAD-WL, Consortium to Establish a Registry for Alzheimer’s Disease Word List Learning Recall Test. AFT, Animal Fluency Test. DSST, Digit Symbol Substitution Test. CI, confidence interval.

Dose-response analyses revealed nonlinear patterns (Figure 3). Delayed memory and verbal fluency exhibited linear declines with increasing exposure duration, whereas working memory (DSST) demonstrated a biphasic response, initial stability followed by deterioration after prolonged exposure (>30 years). A latency window of 15–25 years preceded measurable cognitive impairment, with the most severe effects emerging after 30 years of cumulative exposure.

**Figure 3.**
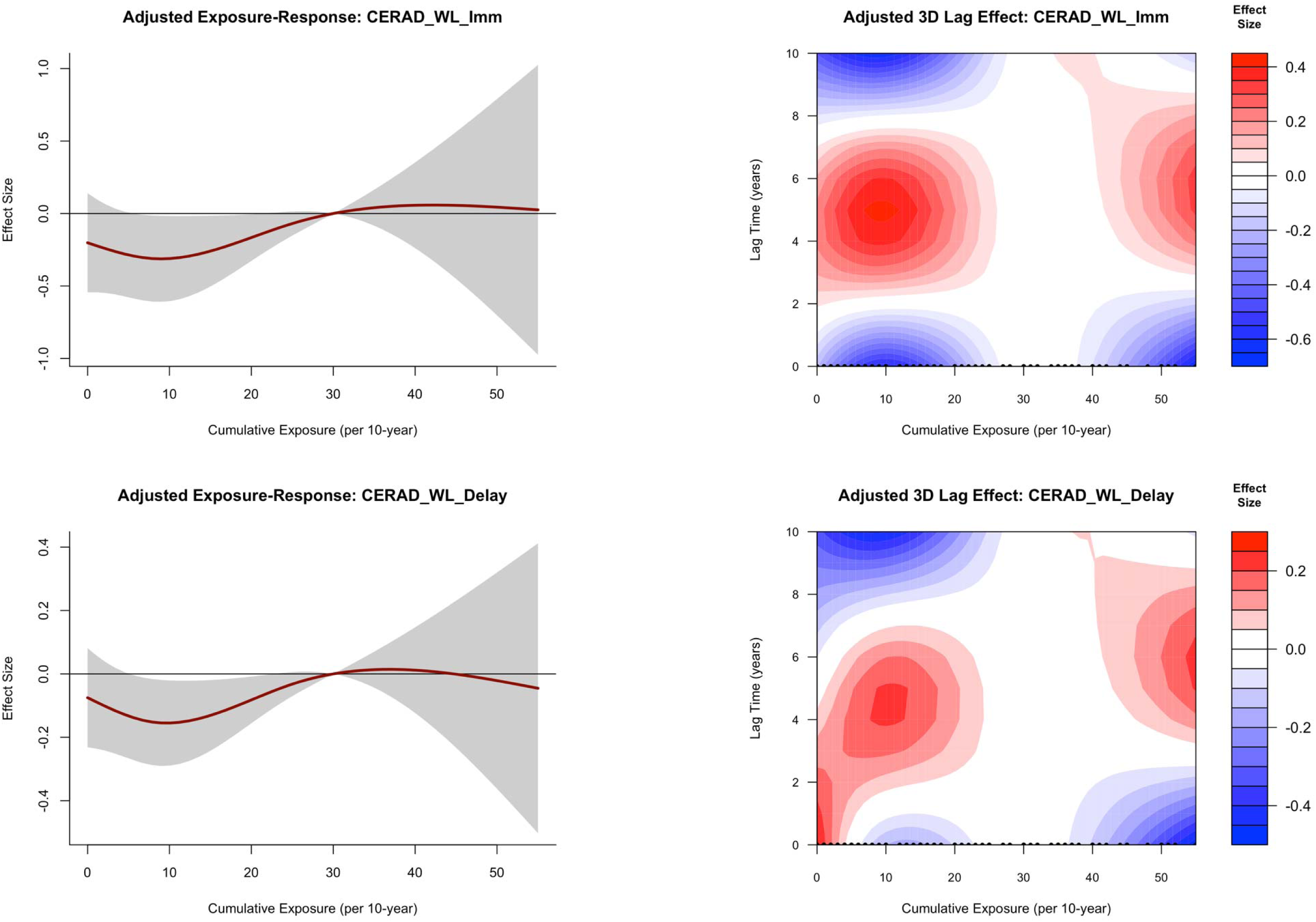
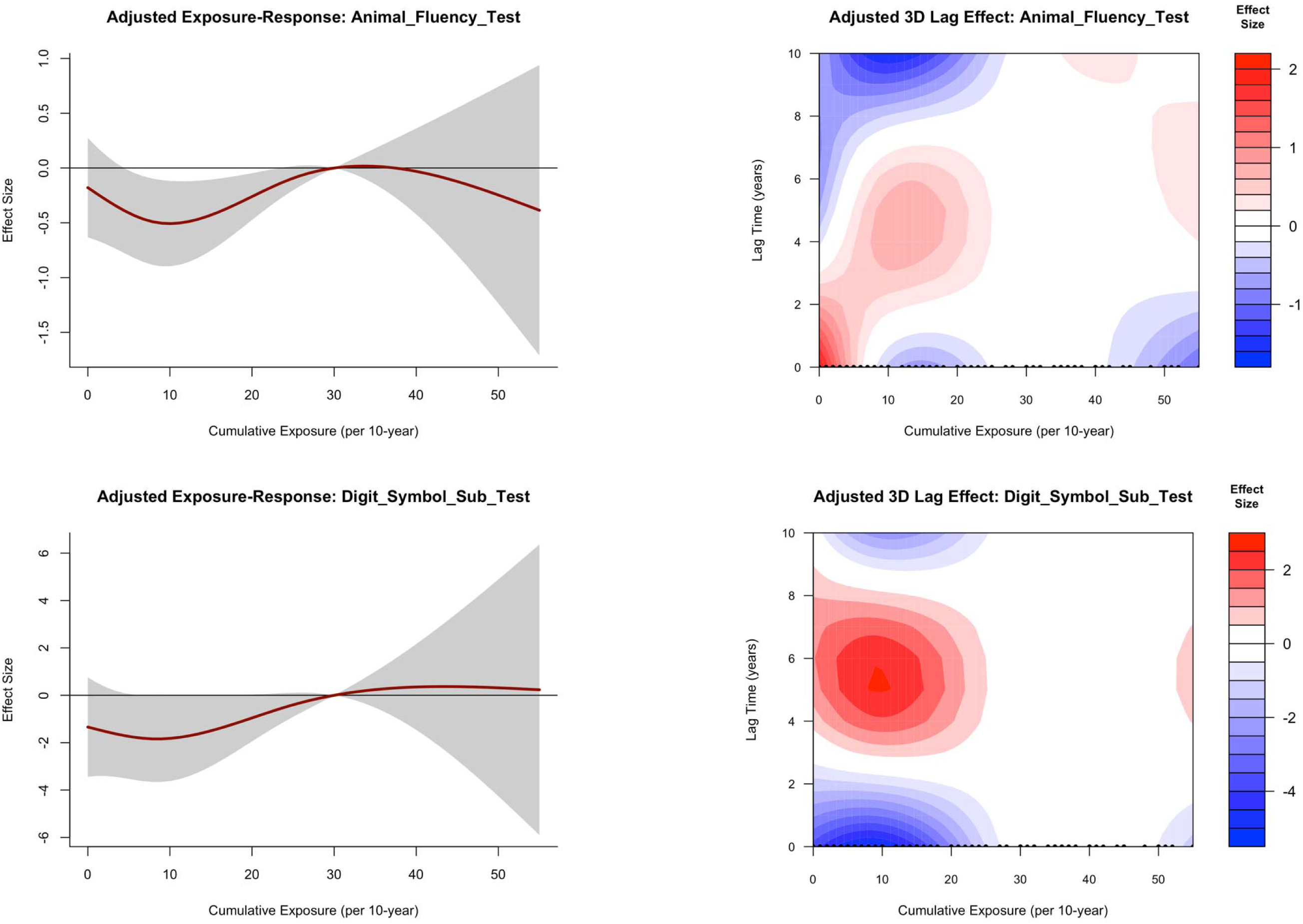
Associations Between Occupational Exhaust Fumes Exposure and Cognitive Decline in NHANES Participants. The exposure-response curves show the adjusted dose-dependent effects of exposure on cognitive outcomes, with the effect sizes quantified as changes in cognitive scores per unit (10 years) increase in exposure. The three-dimensional contour plots show the adjusted joint effects of exposure intensity and duration (lag time) on cognitive function scores, showing biphasic pattens characterized by initial adaptive responses followed by potential neurotoxic effects at higher cumulative exposures. Logistic regression models with DLNMs were adjusted for age, gender, educational attainment, race, history of CVD, depression and diabetes. The solid lines represent the estimated effect sizes from the fitted DLNM, and the shaded areas represent the 95% confidence intervals around the estimate for each exposure level. Notes: NHANES, National Health and Nutrition Examination Survey. CVD, cardiovascular diseases. DLNM, distributed lag nonlinear model.

Analysis of the inter-relationships between cognitive tests revealed that correlations among verbal fluency, memory, and working memory were generally stronger in the exposed group (Supplementary Figure s1). We further visualized the patterns of multi-domain cognitive impairment using a Sankey diagram (Figure 4). This analysis revealed the proportions of individuals with impairments in all three tests, in both AFT and DSST, and with impairment exclusively in CERAD-Delayed Recall were significantly higher in the exposed group.

**Figure 4.**
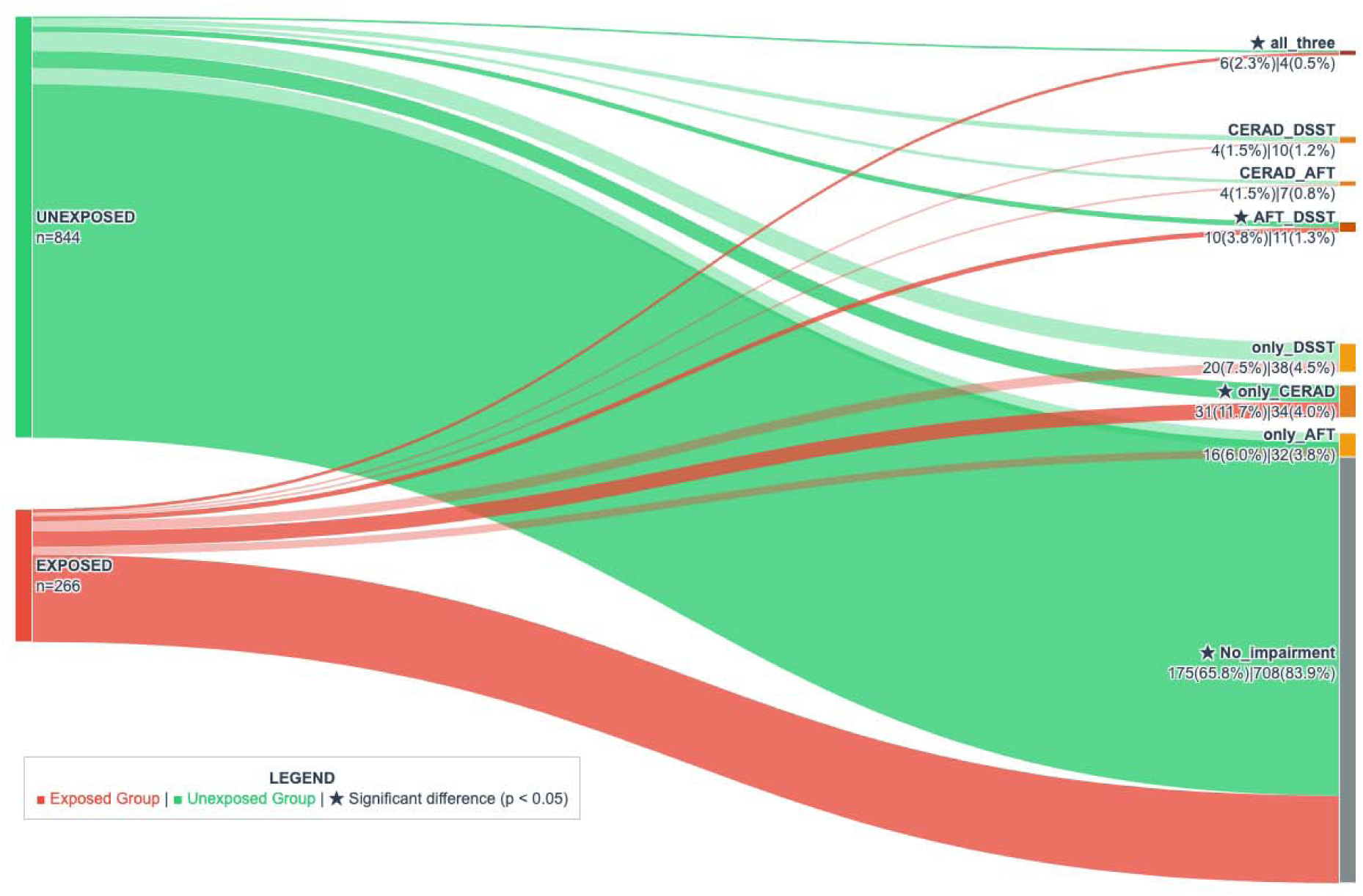
Sankey Diagram Visualizing Patterns of Cognitive Impairment by Occupational Exhaust Fume Exposure Status. In the diagram, the vertical bars from left to right represent the exposure status of participants and their cognitive impairment profiles. Each flow goes from an exposure group (left bar) to a specific pattern of cognitive test results (right bar). The flow width indicates the number of participants within each impairment category. The two bars on the right collectively show the distribution of impairment patterns, ranging from no impairment in any test to impairment in all three tests. Statistical significance (p < 0.05) for between-group differences in specific categories is marked with a star (*). Notes: CERAD, Consortium to Establish a Registry for Alzheimer’s Disease Word List Learning Recall Test (Delayed Recall); AFT, Animal Fluency Test; DSST, Digit Symbol Substitution Test.

### 3.3 Stratified Analyses

In stratified analyses (Supplementary Figure s2), gender differences did not reach statistical significance, though women showed a consistent trend of higher impairment across all four cognitive tests (ORs > 1). Racial differences were substantial, with Mexican American/Other Hispanic and non-Hispanic Black participants showing significantly increased risks in AFT and DSST (ORs up to 4.28), whereas non-Hispanic Asian and other groups showed lower or non-significant risks. Analysis of educational disparities indicated that participants with lower education exhibited a trend toward higher impairment in CERAD-WL Immediate and Delayed Recall (ORs ∼1.26-1.29), though this association was not significant. These findings suggest that race may act as a modifier of the association between occupational exposure and cognitive outcomes.

## 4. Discussion

This study provides novel evidence that occupational exposure to exhaust fumes is associated with domain-specific cognitive decline in older adults. The magnitude of the association is significant, with profound dose-response relationship after 30 years of cumulative exposure. Our findings extend previous research on environmental air pollution by demonstrating that occupational settings health (Carey et al., 2018; Shaffer et al., 2021; Gao et al., 2022), characterized by higher and more sustained exposure levels, present unique risks to cognitive .

The DLNM analyses provided several critical insights into the temporal dynamics of exhaust fume-related neurotoxicity. We identified a latency window of 15-25 years between exposure onset and measurable cognitive decline, with the most severe impairments becoming apparent after 30 years of cumulative exposure. This delayed manifestation parallels findings from studies of other occupational neurotoxicants, including pesticides and industrial dusts (Pedersen et al., 2019; Alif et al., 2023), suggesting a common pattern of neurological damage. The biphasic dose-response relationship we observed - where low-level exposure (<20 years) transiently improved working memory while prolonged exposure (>30 years) led to significant impairment - may reflect initial neuroadaptive responses mediated by Nrf2 pathway activation (Mayer et al., 2024), followed by the eventual breakdown of compensatory mechanisms under sustained oxidative stress (Block et al., 2009; Milani et al. 2018).

The patterns of cognitive impairment, visualized by the Sankey diagram, provide critical behavioral evidence for the progression from compensatory adaptation to systemic failure. While the stronger inter-domain correlations (Supplementary Figure s1) suggest an initial reorganization of cognitive networks, meaning a compensatory integration to maintain function under early neurotoxic stress, the Sankey diagram reveals the ultimate consequence of prolonged exposure: a significantly higher prevalence of co-occurring impairments across multiple cognitive domains. The exposed group exhibited a 4.6-fold higher rate of triple-domain impairment (involving memory, verbal fluency, and processing speed) and a significantly lower proportion of cognitively intact individuals. This shift from isolated to diffuse impairment profiles indicates that the brain’s compensatory capacity becomes overwhelmed, leading to widespread cognitive dysfunction. This trajectory perfectly aligns with the biphasic dose-response observed in our DLNM analyses and mirrors the compensatory failure seen in preclinical neurodegenerative states (Cabeza et al., 2018).

The neurotoxic mechanisms underlying these associations likely involve multiple interacting pathways. Exhaust particles can induce mitochondrial dysfunction via excessive reactive oxygen species production (Zheng et al., 2019; Ihantola et al., 2020), particularly affecting dopaminergic neurons in a manner resembling early Parkinsonian pathology (Andican et al., 2012). Concurrently, these particles trigger neuroinflammatory cascades, with microglial activation and pro-inflammatory cytokine release (e.g., TNF-α, IL-6) contributing to synaptic damage and neuronal loss (Li et al., 2020; Pradhan et al., 2023). Notably, emerging evidence suggests that exhaust fumes may also impair neuronal autophagy independently of microglial activation (Barnhill et al., 2020; Ha et al., 2022), representing a direct pathway for neurodegeneration that may be particularly relevant in occupational settings with high-intensity exposures (Berr et al., 2019).

Our domain-specific findings align well with these mechanistic insights. The early, transient improvement in working memory (mediated by prefrontal circuits) may reflect temporary adaptive responses in brain regions with greater neuroplasticity (Calabrese et al., 2017; Luciana & Collins, 2022), while the later declines in verbal fluency and delayed memory (linked to temporal lobe structures) correspond to areas more vulnerable to cumulative damage (Corriveau-Lecavalier et al., 2024; Wuestefeld et al., 2024). This pattern of regional vulnerability mirrors what is seen in neurodegenerative diseases (Yorifuji et al., 2016), suggesting possible overlaps in pathogenic mechanisms.

Beyond the biological mechanisms, our findings also carry important implications for occupational health policy. The observed association between long-term exhaust fume exposure and domain-specific cognitive decline suggests that current occupational exposure standards may underestimate neurocognitive risks. Strengthening labor protection regulations, such as stricter exposure limits, improved ventilation, and mandatory use of personal protective equipment, could help mitigate these risks. Regular cognitive screening programs for workers with extended exposure histories may facilitate earlier detection and intervention, while workplace health policies should add cognitive function monitoring along with traditional cardiopulmonary surveillance.

Several limitations must be acknowledged. The cross-sectional design precludes definitive causal inferences, and the reliance on self-reported exposure histories introduces potential recall bias. Furthermore, we were unable to account for all potential co-exposures (e.g., noise, other chemicals) that may have contributed to the observed effects (Ren et al., 2024; Martins et al., 2025). The lack of biomarker data also limits our ability to directly link the cognitive changes to specific neuropathological processes. To address this, we will conduct metabolomic analyses within this exposed cohort to characterize metabolite profiles associated with cognitive impairment, with the goal of defining a distinct metabolomic signature indicating exhaust fume-induced cognitive impairment.

## 5. Conclusion

Due to the cross-sectional nature of this study, our findings suggest an association between prolonged occupational exposure to exhaust fumes and cognitive decline in older adults, particularly affecting memory and verbal fluency; however, causality cannot be established. The dose-response relationship, with significant effects emerging after 30 years of exposure, highlights the cumulative neurotoxicity of workplace pollutants. These results underscore the need to incorporate cognitive health endpoints into occupational safety standards and implement targeted protections for high-exposure workers. Future research should prioritize longitudinal studies to establish causality and incorporate biomarker assessments to validate these findings and explore underlying mechanisms.

## Data availability statement

Publicly available datasets were analyzed in this study. This data can be found here: the datasets for this study can be found in the NHANES repository. Please see the https://wwwn.cdc.gov/nchs/nhanes/Default.aspx for more details.

## Ethics statement

The studies involving human participants were reviewed and approved by the Research Ethics Review Board (ERB) of the US National Center for Healthcare Statistics (NCHS) authorized the 2011–2012 NHANES (Protocol Number: Protocol #2011-17). The patients/participants provided their written informed consent to participate in this study. Written informed consent was obtained from the individual(s) for the publication of any potentially identifiable images or data included in this article.

## CRediT authorship contribution statement

Lili Liang: conceptualization, formal analysis, writing original draft, review and editing.

Perry E Sheffield: conceptualization, methodology, editing and review.

Rose Saint Fleur-Calixte: software, and formal analysis.

Tianxu Xia: methodology and review.

Spencer Xinyi Zhang: validation and writing original draft.

Jenny J Lin: methodology, review and editing.

## Acknowledgments

We express our special appreciation to Dr. John Doucette from the Icahn School of Medicine at Mount Sinai for his invaluable support on statistical consultation. We are grateful for the NHANES team and all participants.

## Supplementary materials

Supplementary materials to this article can be found online.

