## Supplemental file for "Long-term Occupational Exhaust Fumes Exposure and Delayed Cognitive Impairment in Older U.S. Adults: A Cross-sectional Study in U.S"

**Table s1. Baseline Characteristics of Participants Who Completed vs. Did Not Complete All Cognitive Tests (n = 1207).** Notes: Educational Attainment: < HSG: less than 12th grade, including 12th grade with no diploma; HSG: high school graduate or GED; >HS: some college or associate degree, and college graduate or higher).

|  |  | **No.**  **(%)** | **Completed group**  **No. (%)** | **Uncompleted group**  **No. (%)** | ***P* Value** |
| --- | --- | --- | --- | --- | --- |
| **Total** |  | **1207**  **(100.0)** | **1110**  **(92.0)** | **97**  **(8.0)** |  |
| **Age (years)** |  |  |  |  | ***0.0253*** |
| 60-69 |  | 779  (64.5) | 727  (60.2) | 52  (4.3) |  |
| 70-79 |  | 428  (35.5) | 383  (31.7) | 45  (3.7) |  |
| **Gender** |  |  |  |  | *0.2713* |
| Female |  | 581  (48.1) | 540  (44.7) | 41  (3.4) |  |
| Male |  | 626  (51.9) | 570  (47.3) | 56  (4.6) |  |
| **Race/Ethnicity** |  |  |  |  | ***0.0001*** |
| Mexican American and Other Hispanic |  | 235  (19.4) | 214  (17.7) | 21  (1.7) |  |
| Non-Hispanic White |  | 451  (37.3) | 435  (36.0) | 16  (1.3) |  |
| Non-Hispanic Black |  | 384  (31.9) | 347  (28.8) | 37  (3.1) |  |
| Non-Hispanic Asian and Other Race |  | 137  (11.4) | 114  (9.5) | 23  (1.9) |  |
| **Education Attainment** |  |  |  |  | ***0.0010*** |
| < HS |  | 332  (27.5) | 290  (24.0) | 42  (3.5) |  |
| HSG |  | 264  (21.9) | 245  (20.3) | 19  (1.6) |  |
| > HS |  | 611  (50.6) | 575  (47.6) | 36  (3.0) |  |
| **Exposure to Exhaust Fumes** |  |  |  |  | *0.8727* |
| Yes |  | 288  (23.9) | 266  (22.1) | 22  (1.8) |  |
| No |  | 919  (76.1) | 844  (69.9) | 75  (6.2) |  |

**Figure s1. Correlation Matrices of Cognitive Test Scores by Exposure Status.** (E): Exposed group (Lower Triangle); Unexposed group (Upper Triangle). Correlation coefficients were calculated using Spearman's rank correlation. Values indicate Spearman's ρ. The correlations among verbal fluency, memory and processing speed were stronger in the exposed group. Notes: CERAD-WL, Consortium to Establish a Registry for Alzheimer’s Disease Word List Learning Recall Test. CERAD-WL Imm, CERAD-WL Immediate Recall. AFT, Animal Fluency Test. DSST, Digit Symbol Substitution Test.

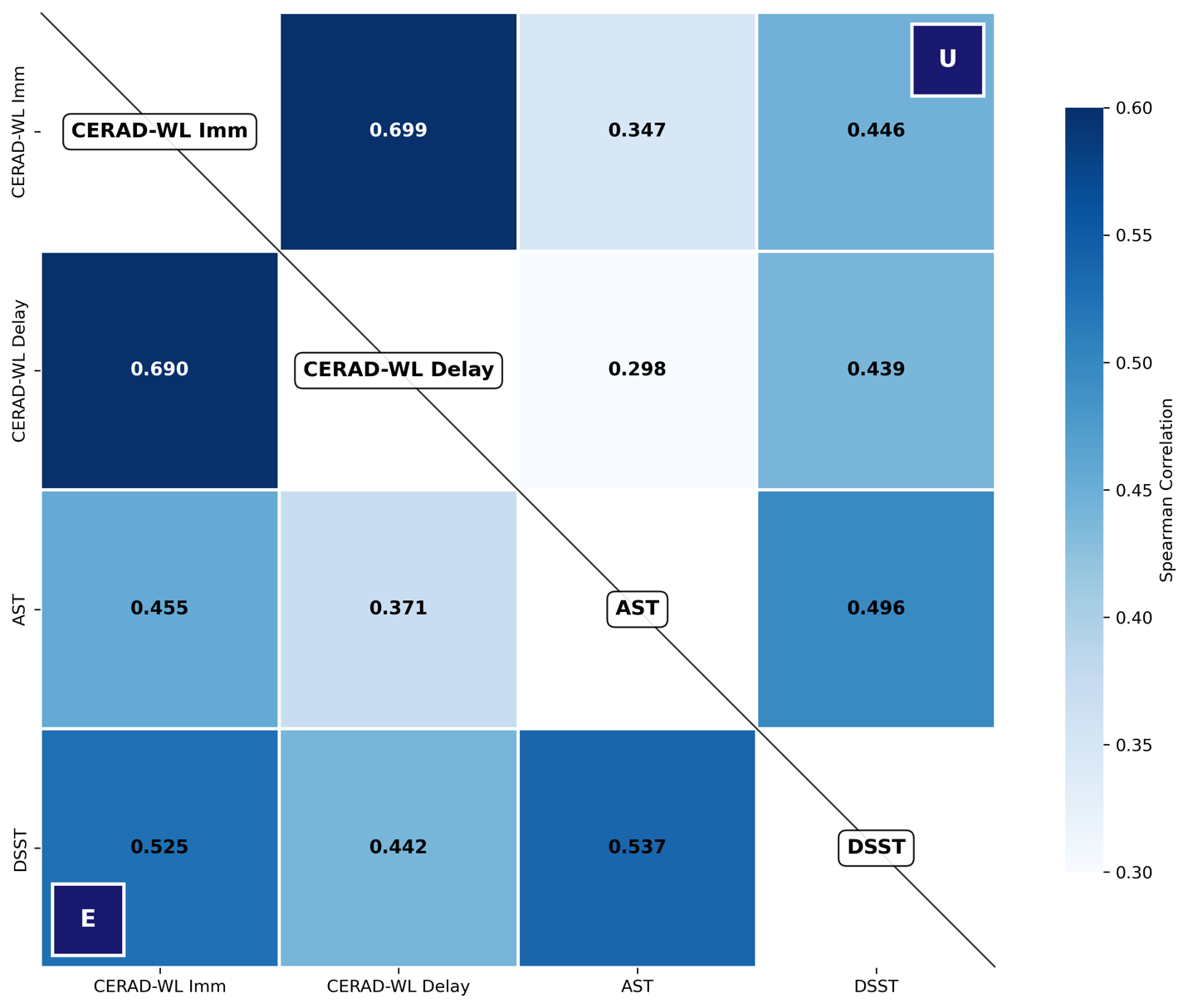

**Figure s2. Differences in Cognitive Impairment Associated with Occupational Exhaust Fume Exposure by Gender, Race, and Educational Attainment.** Forest plots display odds ratios (ORs) with 95% confidence intervals (CIs). An OR > 1 indicates higher impairment risk in each subgroup compared to their reference group: (A) Male; (B) Non-Hispanic White; (C) > HS. Notes: CERAD-WL, Consortium to Establish a Registry for Alzheimer’s Disease Word List Learning Recall Test; CERAD-WL Imm: CERAD-WL Immediate Recall; CERAD-WL Delay: CERAD-WL Delayed Recall; AFT, Animal Fluency Test; DSST, Digit Symbol Substitution Test. Educational Attainment: < HSG: less than 12th grade, including 12th grade with no diploma; HSG: high school graduate or GED; >HS: some college or associate degree, and college graduate or higher).

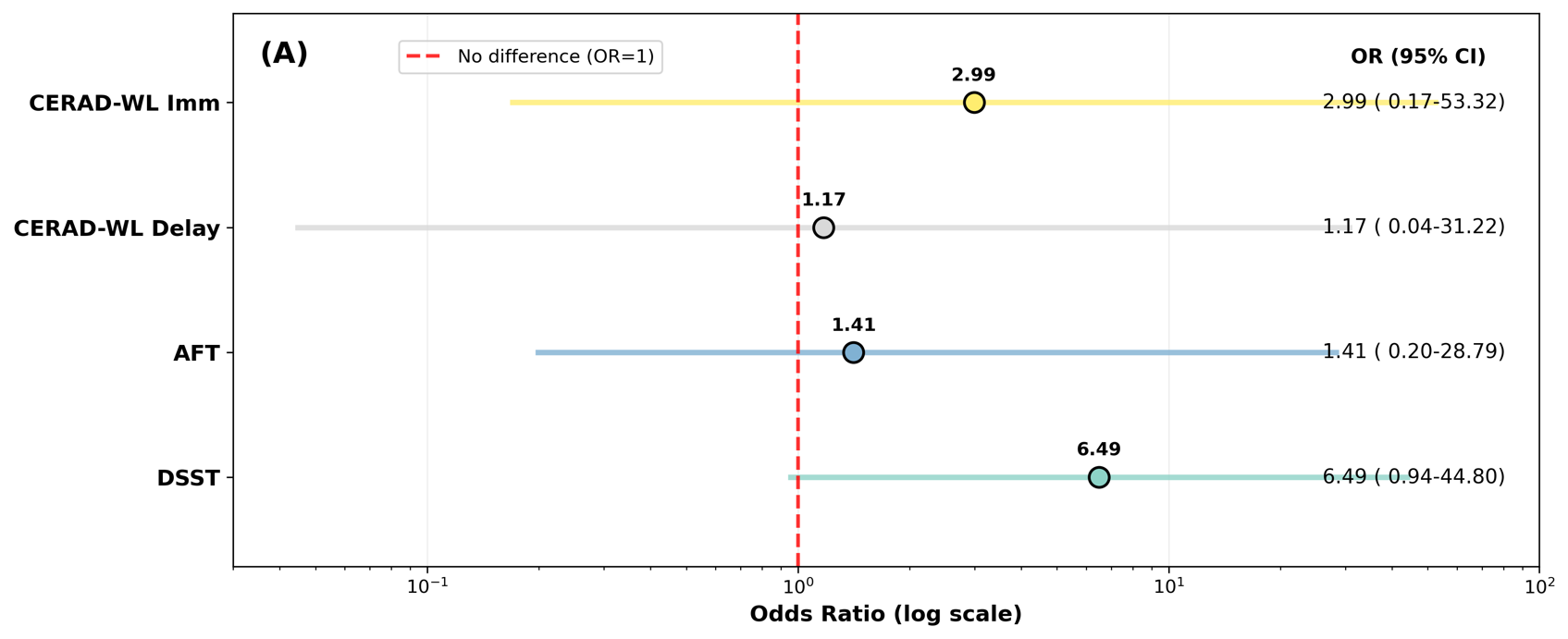

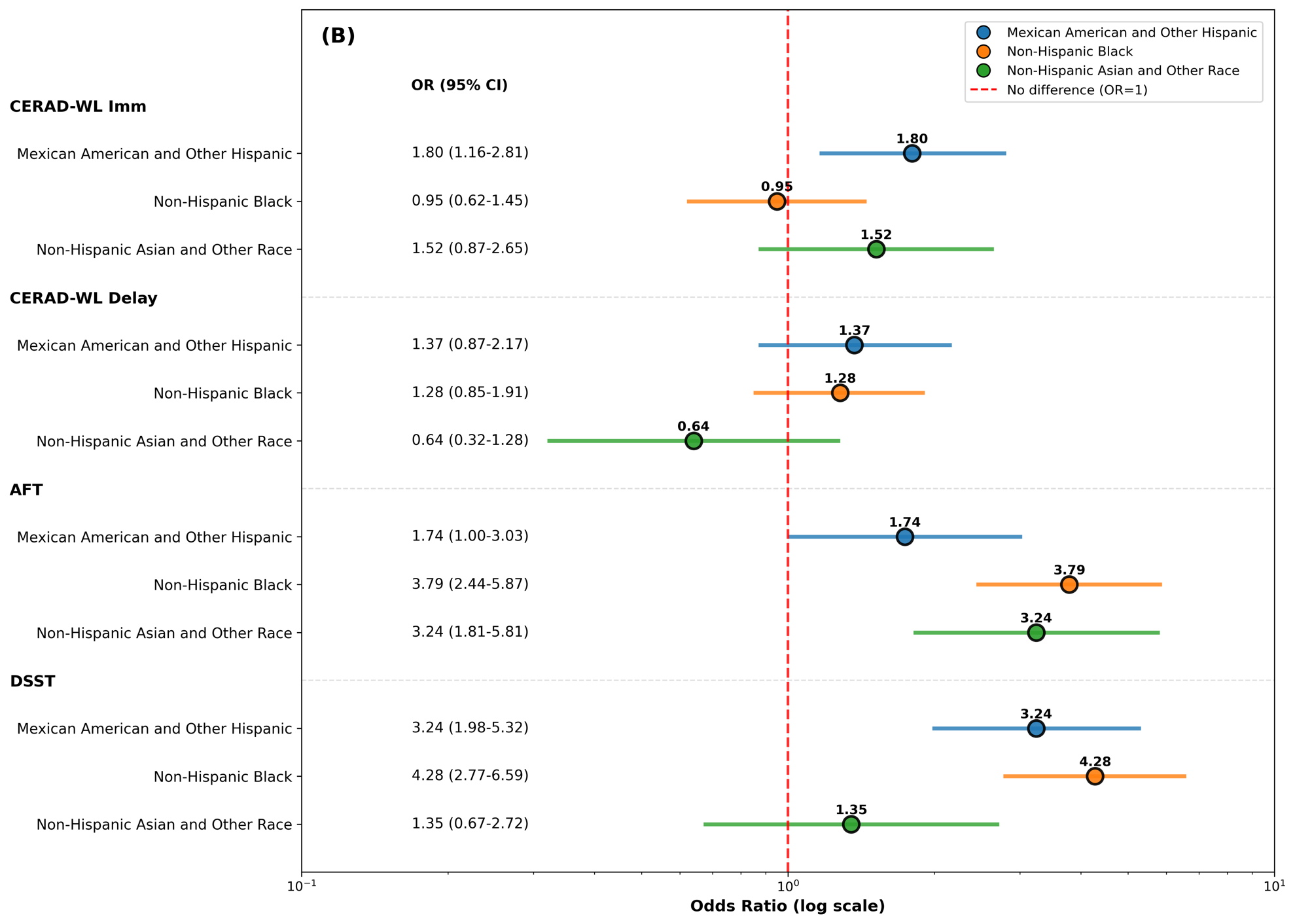

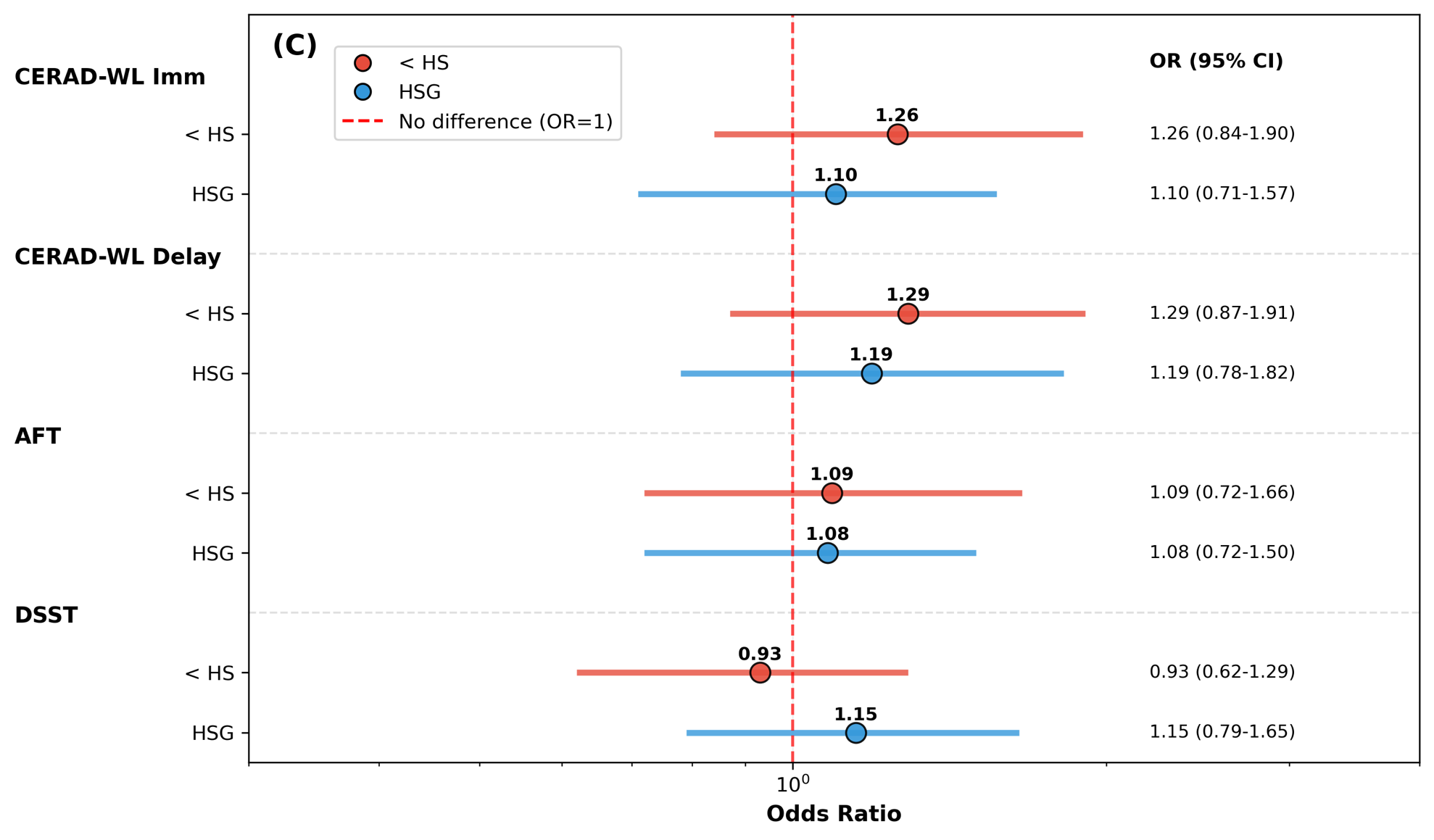
